# Known Group Validity of Assessment of Work Performance for Thai Homeless People

**DOI:** 10.1101/2025.01.23.25321060

**Authors:** Uthaikan Thanapet, Watthanaree Ammawat, Maliwan Rueankam, Winai Chatthong, Supalak Khemthong

## Abstract

**Background:** Substance use and mental disorders are highly prevalent in Thai homeless people living with no mental health recovery. The lack of psychosocial occupational therapists may be one huge problem for none of the measurement outcomes for clinical utility. We aimed to determine the known-group validity of the Assessment of Work Performance (AWP) with good content and internal consistency reliability.

**Methods:** Participants (N = 60) were recruited by screening with no cognitive impairment, no psychiatric symptoms, independent self-care, and on-the-job assignment at Nonthaburi’s destitute home. Mann–Whitney U tests firstly found a significant difference in process skills between males and females (p < .05). Data analysis of the ceiling effect was then conducted to design using cleaned samples (N = 22), and the Mann–Whitney U tests secondly found a significant difference in communication skills between males and females (p < .05).

**Results:** There were no associations between the frequency of genders and substance abusers were computed using cross-tabulation (N = 60 versus N = 22). Besides a corrected sample, natural contexts of too easy job selection, no preparatory of supported employment, and non-purposeful living activities may be confounding factors of avolition, poor habit, and inactive life roles without wellness at the end of life.

**Conclusion:** These preliminary results suggest a useful AWP with good known-group validity for measuring outcomes among vulnerable people and describing how to improve motor, process, and interactive communication skills based on the Model of Human Occupation (MOHO) in occupational therapists and Mental Health Recovery (MHR) workers.

## Background

A personal belief about how they can achieve sensing of hope based on Mental Health Recovery or MHR [1] for instance, productive services and positive motivation are promoted by having progressive self-enhancement [2] with the goal-directed level of cognitive, optimistic socialization, and in-person physical and psychosocial rehabilitation for promoting a socially inclusive support system of supported employment. As shown in the duration of 6-year follow-up in people with serious mental illness (SMI), cognitive flexibility is prioritized as a key outcome within an existing service delivery model [3].

This outcome is significantly associated with community functioning [3]. Interestingly, the second upskilling of a collaborative learning model via recovery colleges [4] is demonstrated to improve innovatively education-based MHR approach. Hopkins [5] and Khemthong [6] have also mentioned that recovery colleges offer safe environment-related social opportunities and supportive choices to be a new hub of learning new knowledge and skillful caring interaction for further structuring day-time activities. Jasni and colleagues [7] explored knowledge of government caregivers’ experiences with homeless former prisoners in Malaysia. Triangulation found that internal migration causes a high prevalence of homelessness in an urban population, leading to limited employment opportunities and residential areas. Consequently, Honey et al. [8] interviewed 61 people who accessed a new MHR service and were interviewed about their experiences, including how the service had influenced their sense of hope. Using constant comparative analysis found a sense of hope improved positive changes in themselves with new understandings to manage challenges by having progressive plans and supportive services of accessibility, staff competence (wisdom), and caring interactions.

Nous Group × The University of Sydney [9] used a qualitative content analysis of data among people who had accessed HeadtoHelp, at one state of 15 new MHR services. One-to-one talking therapies were provided by nurse practitioners, occupational therapists, psychiatrists, and peer workers. However, the program evaluation showed inappropriate numbers, difficulties with staff recruitment, and unclear identifying the types of services that require assessment tools to meet clients’ demands. For instance, an assessment of work performance or AWP is the first observation-based assessment of personal abilities including physical, process, and communication/integration skills [10, 11].

The AWP was created by Sandqvist et al. [11] to assess personal efficiency under structured circumstances using real work-life tasks in a wide range of disability [12] e.g. Turkish people with intellectual disability [13], Swedish people in either physical or mental health problems [10]. However, no study of the AWP has been found to assess Thai homeless people’s performance in terms of motor, process, and interactive skills while working on duties. Therefore, this pilot study will bring a crucial tool after testing known-group validity to Thai occupational therapists and MHR workers to enable work performance by revealing their possible explanations based on the Mode l of Human Occupation (MOHO).

## Materials and methods

### Study design

This cross-sectional test-retest reliability study was conducted. Interprofessional collaboration was communicated among social workers, psychologists, occupational therapy students, and academics. A permission agreement contract was made to translate the AWP to Thai (version 2.0) between Unitalent representing Jan Sandqvist and the corresponding author on 15 Dec 2022. This study has received ethical approval from the Mahidol University Central Institutional Review Board (COE No. MU-CIRB 2023/186.2212). Permission to use consent information and assessment tools was considered for safety and confidentiality by the Department of Social Development and Welfare, Ministry of Social Development and Human Security, THAILAND.

### Participants and data collection

Purposive samplings were used among 212 homeless people who were classified independent level of self-care abilities living at Nonthaburi’s destitute home. The interview protocol contains three parts. Part A included questions on the socio-demographics of the participants. Part B comprised a cut-off equal to or more than 23 scores of the Rowland Universal Dementia Assessment (RUDAS) and a cut-off equal to or less than 36 scores of the Brief Psychiatric Rating Scale (BPRS). Part C consisted of a checklist of inclusion criteria: (1) be over 18 years old and no later than 64 years, (2) born in Thailand and used the Thai language; (3) be diagnosed with mental disorders – no delusion and acute episode, (4) have no physical handicap, (5) Able to use both hands – no neurological deficits, and (6) have no low vision/deafness. All sample sizes (n = 60) have been calculated using the statistical power at 80% with a confidential level of 95% to protect against bias. Sixty participants were finally selected out of the 72 participants.

### Assessment tools

A demographic form was used in this study including age, gender, diagnosis, experienced drug addiction, living duration, and received non-chronic disease/s. The AWP (Thai version) was also implemented after its forward-backward translation and content validity was acceptable agreement. Although a specific task was not required for the AWP, those homeless participants continued undergoing the virtual reality (VR) programs: two-month supported employment and in-house pre-vocational training. The AWP was designed to assess three skills of work performance: 5 items of motor skills, 5 items of process skills, and 4 items of communication/interaction skills [11]. Quality of work performance was observed and noted in terms of efficiency, adequacy, and appropriateness on its blank box. Scoring was rated per one item using a four-point ordinal scale (1 = deficient performance, 2 = inefficient performance, 3 = uncertain performance, 4 = competent performance). If an assessor did not know ade quate information or relevant situations to rate each item in the client’s working context, the alternative notes would be lack of information (LI) and not relevant (NR) respectively. The total administration time of rating the 14-item AWP was a flexible observation, depending upon the working situation of individuals. When the assessors noted LI or NR, the summation of the total score could not be completely computed [10, 12].

The corresponding author translated an original English version of AWP into Thai. Then, a bilingual person translated the Thai version into English (the 2^nd^ draft). After that Jan Sandqvist approved the Thai version 2.0 of AWP on 29 August 2023. The content validity index, or CVI was accepted gaining 1.00 for all items from thr ee experts in psychosocial occupational therapy. Some useful comments from those experts were brought to correct all keywords becoming the Thai version 3.0 of AWP which was applied to those participants (n = 60). Consequently, the researchers asked one psychologist and one social worker for appropriate tasks as a job selection to be observed in the AWP. The existing two-month VR programs were selected: supported employment as community gardening and in-house pre-vocational training i.e., rug hooking in a tutorial group, cleaning tables in a food corner, and sweeping the leaves/growing small plants. Six occupational therapy students were assigned to observe and score those participants while doing the VR programs. The 14-item AWP Thai version was tested gaining an acceptable Cronbach’s alpha of 0.68 [14].

### Statistical analysis

Statistically, IBM SPSS V.22 was used for testing known-group validity. This type of validity is appropriate when there is no comparison of a gold standard. Additional variables of interest included gender and substance abuse experience. Mann–Whitney U tests were conducted to determine if differences exist on the ordinal scales [15]. Flooring and ceiling effects were also investigated calculating the number of people who performed in the lower and upper 10% of the total score [16]. The chi-square test was finally used to determine frequencies in a 2*2 cross-tabulation of those additional variables [15].

## Results

Demographically, this study included male subjects (63.3%) highly outnumbered female subjects (36.7%). Almost half the subjects (38.3%) graduated from primary school, never went to school (6.7%), kindergarten (13.3%), secondary (15%), high school (16.7%), and high vocational school (8.3%). Only 1.7% completed a university degree. The participants’ mean age was 44.10 years, standard deviation (SD) 7.68. They lived at Nonthaburi’s destitute home for an average of 53.18 months, SD 37.68. A majority (63%) had unspecified mental disorders diagnosed; of those, 35.31% had schizophrenic spectrum, and 1.69% had mood disorders. Over half (55%) of the sample had experienced substance abuse.

In Table 2 Mann–Whitney U tests indicated no significant difference between male and female participants for the mean of motor domain, interactive communication domain, and total score on the AWP (p > .05). Mann–Whitney U tests indicated significantly higher scores of process skills for male (n = 38) than female (n = 22) participants (Z = −2.519, p = .012). All three domains and the total score on the AWP for the participants who had experienced substance abuse (n = 33) were compared to those with no experience (n = 27). Mann–Whitney U tests indicated no significant difference (p > 0.05).

**Table 1.** Demographic percentage of the participants (n = 60)

| Demographic data | Percentage | Mean (SD) |
| --- | --- | --- |
| **Age (years)** |  | 44.10 (7.68) |
| **Living duration (months)** |  | 53.18 (37.68) |
| **Gender** |  |  |
| male | 63.3% |  |
| female | 36.7% |  |
| **Education** |  |  |
| never went to school | 6.7% |  |
| kindergarten school | 13.3% |  |
| primary school | 38.3% |  |
| secondary school | 15.0% |  |
| high school | 16.7% |  |
| high vocational school | 8.3% |  |
| university/undergraduate | 1.7% |  |
| **Diagnosis** |  |  |
| schizophrenia, schizotypal, & delusional disorders | 35.31% |  |
| mood disorders | 1.69% |  |
| unspecified mental disorders | 63.00% |  |
| **Had experienced substance abuse** | 55.00% |  |

**Table 2.** Mann–Whitney U tests between genders and substance abusers (N = 60)

| AWP domains | Mean (SD) |  |  |  |
| --- | --- | --- | --- | --- |
|  | Male<br>n = 38 | Female<br>n = 22 | Substance abuser<br>n = 33 | Not substance abuser<br>n = 27 |
| Motor skills | 19.50 (0.92) | 19.73 (0.63) | 19.61 (0.90) | 19.56 (0.75) |
| Process skills | 19.13 (1.02) | 17.73 (2.66)* | 18.67 (2.01) | 18.56 (1.80) |
| Communication & interaction skills | 14.74 (1.48) | 14.09 (1.85) | 14.70 (1.51) | 14.26 (1.79) |
| Overall skills | 53.37 (2.26) | 51.41 (4.26) | 52.88 (3.33) | 52.37 (3.19) |

\*p < .05; SD standard deviation; AWP, assessment of work performance

The upper 10% of the total score of the AWP was determined for all domains to identify how many participants would have the ceiling effect. As seen in Table 4 mean and SD without the ceiling effect were calculated at the corrected range of scores: motor skills (2-18), process skills (2-18), interactive communication skills (1.6-14.4), and overall skills (5.6-50.4). In contrast to the mixed participants (n = 60), the number of participants who scored with the ceiling effect was changed: motor skills (n = 52), process skills (n = 41), interactive communication skills (n = 38), and overall skills (n = 48).

A Mann-Whitney U test was performed again to evaluate whether any significant difference among the number of participants who scored without the ceiling effect. Only one AWP domain (n = 22) indicated that male samples (n = 13) had significantly greater interactive communication skills than female (n = 9) samples, z = [-2.716], p = [.007]. Whereas no significant difference (p > .05) was found in the rest of the AWP domains and total scores, in both comparison of gender and substance abuse experience.

**Table 3.** Number of participants across mixed, corrected × 10% ceiling-scored AWP.

| AWP domains | Mean (SD) |  |  |
| --- | --- | --- | --- |
|  | Mixed | Corrected | Ceiling Scored |
| Motor skills | 19.58 (0.83)<br>n = 60 | 17.75 (0.71)<br>n = 8 | 19.87 (0.34)<br>n = 52 |
| Process skills | 18.62 (1.91)<br>n = 60 | 16.63 (2.29)<br>n = 19 | 19.54 (0.50)<br>n = 41 |
| Communication & interaction skills | 14.50 (1.64)<br>n = 60 | 12.77 (1.48)<br>n = 22 | 15.50 (0.51)<br>n = 38 |
| Overall skills | 52.65 (3.25)<br>n = 60 | 47.33 (3.03)<br>n = 12 | 53.93 (1.45)<br>n = 48 |

SD standard deviation; AWP, assessment of work performance

**Table 4.**
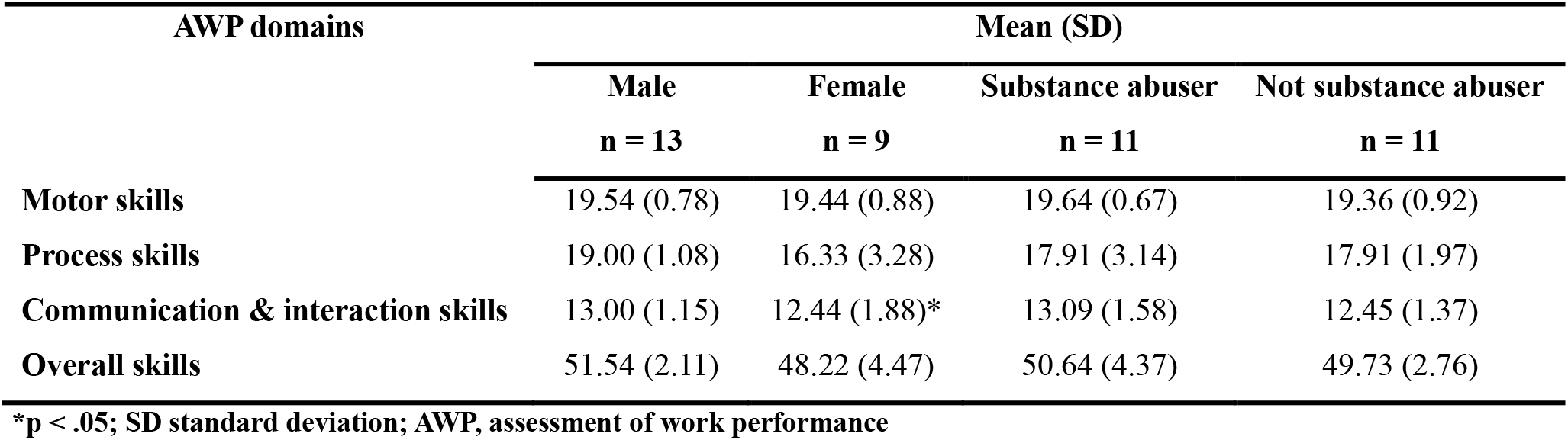
Mann–Whitney U tests between genders and substance abusers (N = 22)

| AWP domains | Mean (SD) |  |  |  |
| --- | --- | --- | --- | --- |
|  | Male<br>n = 13 | Female<br>n = 9 | Substance abuser<br>n = 11 | Not substance abuser<br>n = 11 |
| Motor skills | 19.54 (0.78) | 19.44 (0.88) | 19.64 (0.67) | 19.36 (0.92) |
| Process skills | 19.00 (1.08) | 16.33 (3.28) | 17.91 (3.14) | 17.91 (1.97) |
| Communication & interaction skills | 13.00 (1.15) | 12.44 (1.88)* | 13.09 (1.58) | 12.45 (1.37) |
| Overall skills | 51.54 (2.11) | 48.22 (4.47) | 50.64 (4.37) | 49.73 (2.76) |

\*p < .05; SD standard deviation; AWP, assessment of work performance

A chi-square test of independence was also performed in Table 5 to examine the relationship between genders and substance abusers. There was no significant association between those variables, χ^2^(1, N = 60) = 1.279, p = .258; χ ^2^ (1, N = 22) = 0.188, p = .665. However, an expected count of less than 5 was 50% or 2 cells. Thus, this cross-tabulation required a Fisher’s exact test instead for p = .500 if over 20% of the expected count of less than 5.

**Table 5.**
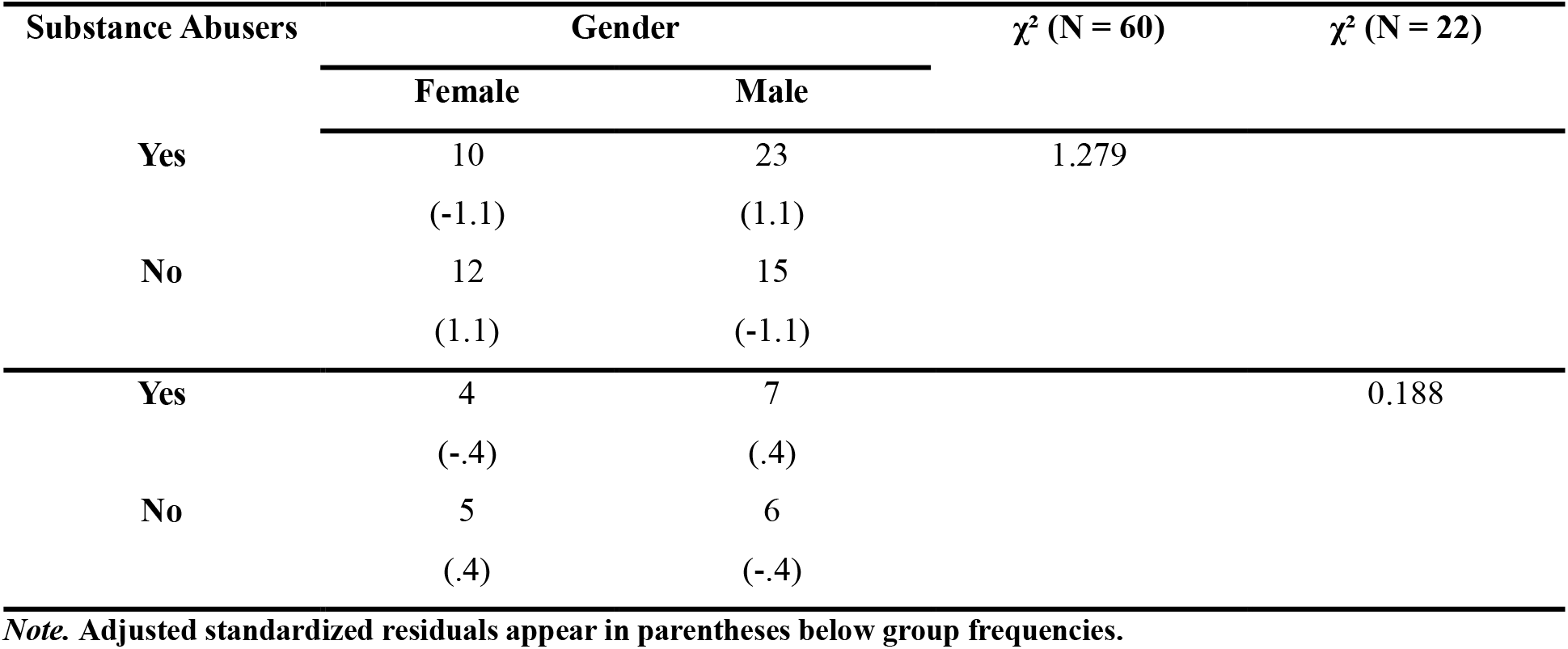
Cross-tabulation results between genders and substance abusers.

| Substance Abusers | Gender | | $\chi^2$ (N = 60) | $\chi^2$ (N = 22) | |
| --- | --- | --- | --- | --- | --- |
|  | Female | Male |  |  |  |
| Yes | 10 | 23 | 1.279 |  |  |
|  | (-1.1) | (1.1) |  |  |  |
| No | 12 | 15 |  |  |  |
|  | (1.1) | (-1.1) |  |  |  |
| Yes | 4 | 7 |  | 0.188 |  |
|  | (-.4) | (.4) |  |  |  |
| No | 5 | 6 |  |  |  |
|  | (.4) | (-.4) |  |  |  |

*Note.* Adjusted standardized residuals appear in parentheses below group frequencies.

## Discussion

The current study aims to describe the known-groups validity of the AWP implemented in participants (n = 60) who had living experiences with homeless and mental disorders for around 4 years. The researchers hypothesize that known-groups validity would be demonstrated based on group comparisons including gender and substance abuse. Scores on the AWP were not significantly different between gender and substance abuse i n motor, interactive communication, and overall skills when all data was analyzed accounting for the number of participants with and without ceiling effect. Interestingly, the AWP shows a significantly higher score of process skills in the male than female participants (p < .05), in both purposively recruited and specifically corrected data. This result could be based on the complementary roles of occupational therapists in facilitating a hopeful process of MHR experiences [1] for stakeholders at a destitute home through the provision of the AWP as a self-enhancement of belief of community functioning [2, 3, 8].

All participants with mental disorders, either inclusive criteria or ceiling effect correction, are a result of the purposive sampling from interprofessional collaboration focused on the Thai version of AWP. This is a preliminary study regarding vulnerable samples who would attend the behavioral observation-based measurement. Consistent with our hypothesis, scores on the AWP were significantly different for male compared to female participants. Surprisingly, there were no significantly different scores on the AWP between individuals who had experienced substance abuse and no experience. Furthermore, occupational therapists often collaborate to identify preparatory alternatives based on their extensive and shared knowledge of psychosocial and cognitive rehabilitation. The process skills scores on the AWP were higher for male samples than females (p < .05) when the total number of 60 was calculated. In contrast to the total number of samples (n = 22) was based on no ceiling effect, the AWP – interactive communication skills scores were significantly computed. The males significantly performed social components while doing tasks better than the females (p < .05). Finally, the number of participants does not show a critical matter since no association of frequencies demonstrates a finding on cross-tabulation of genders and substance abusers.

Besides the significant scores, the similarity of total AWP scores may be either males or substance abusers who tend to have higher scores than others. This statement reflects the natural circumstance of data collection by observing personal preferences in doing purposeful tasks inside and outside Nonthaburi’s destitute home. This pilot study may have no challenging job to be graded into multitasking since all participants can choose only two choices of work engagement based on gender duties and caregivers’ assignments. Supportive choices of work engagement will alternatively be structured working activities toward learning environments [5, 6]. Theoretically, the AWP provides a reasonable explanation of the Model of Human Occupation (MOHO) regarding how to build performance capacity concerning the activity demands of individuals. To support a work environment, all assessors should be trained by communicating how to use the AWP to evaluate a personal ability to perform tasks and duties effectively and to reflect positive results to improve work performance in actual context [17] by gathering more information on previous working experience [10]. This collaborative learning process seems to be an innovative knowledge asset for MHR colleges [4].

However, the ceiling effect in this current study might be due to the natural problems of participants [10]. It is noteworthy to confirm that the known-group validity of the AWP provides inadequate targeting of highly skilled participants in the motor, process, and interactive communication skills subdomain without ceiling effect. Especially for the physical component, only 8 participants may be questionable which Davutoğlu et al. [13] suggested no less than 10 clients to be calculated per one domain. On the other hand, homeless people with mental disorders might have additional consequences e.g., loneliness, avolition, apraxia, anhedonia, etc. Graded activity and job analysis may be one practical solution to reduce the ceiling effect of the AWP. This statement indicates that volitional assessment in a focused group of different genders should be conducted in parallel to prioritize productive and valuable works before the clinical utility of the AWP on-the-job training as a part of a pre-vocational program leading to supported employment. Further expansion of subdomains, with deficient to inefficient performance, should be considered before job coaching for individuals [10]. Therefore, the significant results of the process and interactive communication skills subdomain indicate a requirement for comprehensive training on the participatory types of observable AWP because of an impairment of functional and social cognition among homeless people with mental disorders.

## Limitations and future direction

A limitation of the current study’s results is that all 14-item AWP cannot be generalized to homeless people with mental disorders across all destitute homes in Thailand. An opportunity for further process is to do a job selection analysis before implementation of the AWP. To prevent floor or ceiling effects under three subdomains, relevant items per one simulated or real-life activity could also be recalculated for acceptable internal consistency reliability whenever new multitasking in one job is introduced to the AWP Thai version for evaluating vulnerable persons. Further investigation of the concurrent validity with other gold standardized tools is advised. Inter-rater reliability of the AWP is also required for other healthcare service providers and caregivers. A good example is a knowledge exploration of Malay caregivers whether they had been experiencing with homeless former prisoners [7].

The second limitation is that the findings may not be generalized to the population due to the nature of its convenience sample. Based on the MOHO, occupational therapists are skilled at assessing the overall performance capability of clients concerning the opened system of living activity environment. As such, occupational therapists must know the teaching of the MOHO and the learning process of how to use AWP to evaluate a client’s ability to perform a wider range of work performance including clients’ workplaces, length of work in current position, and motivative salary, and available job coaching, supportive job agency. Finally, one big limitation is that six occupational therapy students administered the AWP based on the guidelines, the unequal quality of raters may have interfered with the AWP. Useful advice in future studies should design thinking of the AWP online during shelter work at the MHR college. For example, a qualitative content analysis of one-to-one talking therapists [9] and a service delivery model with a 6-year follow-up of cognitive flexibility in people with SMI [3].

## Conclusion

The current study is an important first step to providing a known-group validity scale to assess motor, process, and interactive communication skills for homeless people with mental disorders. A strength of this study is its novelty of four structured activities as a job selection process before data collection of the AWP. Other ongoing improvements include the relatively large sample size and geographically diverse sample of homeless people with mental disorders. The AWP Thai version is a valid tool for assessing personal skills in a vulnerable group, and its internal consistency is further examined to ensure the acceptable Cronbach’s alpha level with new multitasking preferences on personal demands at other destitute homes toward rehabilitation centers.

## Data Availability

All data produced in the present study are available upon reasonable request to the authors

## Abbreviations

AWP: Assessment of Work Performance
MOHO: Model of Human Occupation
MHR: Mental Health Recovery
SMI: Serious Mental Illness
RUDAS: Rowland Universal Dementia Assessment
BPRS: Brief Psychiatric Rating Scale
LI: lack of information
NR: not relevant
VR: virtual reality

## Acknowledgments

The authors would thank occupational therapy students, academics, and experts. They would also like to thank Unitalent representing Jan Sandqvist for their permission agreement for the AWP translation and research. This project would not have been completed without support from all caregivers and clients at Nonthaburi’s destitute home.

## Author contributions

All authors contributed to the study conception and design of the study. Material preparation and data collection was performed by W.C., U.T, W.A and M.R. W.A and U.T had formal analysis. Project administration and data curation were M.R. The first draft of the manuscript was written by S.K and all authors commented on previous versions of the manuscript. S.K and W.C had the main responsibility for writing and revising the manuscript. All authors read and approved the final manuscript.

## Declarations

### Ethics approval and consent to participate

This study has received ethical approval from the Mahidol University Central Institutional Review Board (COE No. MU-CIRB 2023/186.2212). Permission to use consent information and assessment tools was considered for safety and confidentiality by the Department of Social Development and Welfare, Ministry of Social Development and Human Security, THAILAND. Participants were fully informed and provided written consent to participate before conducting the assessment.

### Clinical trial number

TCTR20241122002 https://www.thaiclinicaltrials.org/show/TCTR20241122002 Registration date: 22 November 2024 with Thai Clinical Trials Registry

### Consent for publication

Not applicable.

